# Creatine/creatinine ratio and myostatin as biomarkers to monitor muscle function in Duchenne Muscular Dystrophy patients

**DOI:** 10.1101/2025.08.11.25333307

**Authors:** Chiara Degan, Roula Tsonaka, Sharon I. de Vries, Nadine Ikelaar, Menno van der Holst, Hermien E. Kan, Erik H. Niks, Pietro Spitali

## Abstract

**Objective:** Duchenne Muscular Dystrophy (DMD) is characterized by progressive muscle wasting leading to early loss of motor function. Functional tests monitor disease progression and serve as clinical trial endpoints, but are influenced by maturation in younger patients, variability, and patient motivation. Blood biomarkers, that can predict disease progression and objectively evaluate treatment responses, offer a valuable alternative.

In this study, we investigated whether longitudinal observations of biomarkers myostatin and the creatine/creatinine ratio (Cr/Crn) are associated with functional tests, such as 6-minute walk test, North Star Ambulatory Assessment (NSAA), 10-meter walk-run test velocity, Performance of Upper Limb (PUL2.0), and disease milestones like loss of ambulation (LoA), overhead reach. and hand-to-mouth function.

**Methods:** We used real-world longitudinal data from 74 DMD patients followed for up to 11 years with annual visits to the LUMC outpatient clinic, linked to 408 serum samples. Associations between biomarkers, functional tests, and clinical milestones were assessed using linear mixed models and time-dependent Cox models, respectively.

**Results:** Lower Cr/Crn levels and higher myostatin levels were associated with better functional performance and a less rapid decline in ambulation, given fixed treatment and BMI. Children with one-unit higher log2-myostatin levels had, on average, 4.73 points higher NSAA and 3.40 points higher PUL2.0 (both p < 0.001), and were 42% less likely to lose ambulation over the following year. Conversely, children with one-unit lower log2-ratio levels had, on average, 7.18 points higher NSAA and 11.40 points higher PUL2.0 (both p < 0.001), and were 3.67 times more likely to remain ambulant. We also proved that incorporating log2-myostatin and log2-Cr/Crn as endpoints could reduce the required sample size for clinical trials by more than half without compromising statistical power. For instance, to detect a yearly drop of 3 points in the NSAA with 80% power, recruitment requires almost 80 participants in a 1:1 randomized trial, in contrast to a little more than 50 patients for the respective value of log2-myostatin or log2-Cr/Crn.

**Interpretation:** These findings support the potential of myostatin and Cr/Crn as prognostic biomarkers to enhance trial design and endpoint in clinical and interventional trials for DMD.

## Introduction

Duchenne Muscular Dystrophy (DMD) is a rare neuromuscular disorder primarily affecting males, with an incidence of less than 1 in 5000 males and fewer than 1 per million females^1^. It is caused by a mutation in the *DMD* gene, which leads to delayed motor development due to progressive loss of muscle mass and muscle function, early loss of ambulation, and reduced life expectancy. As the disease advances, individuals become increasingly dependent on assistive devices, often requiring wheelchairs and ventilatory support, and may develop cardiac and respiratory complications.

Functional tests, such as 6-minute walk test (6MWT), North Star Ambulatory Assessment (NSAA), 10-meter walk-run test velocity (10MWT), and Performance of Upper Limb (PUL2.0), are used to evaluate disease progression in routine clinical visits and serve as clinical trial endpoints^2–4^. However, these tests are often affected by patient motivation, particularly in young children, and the evaluator’s ability to effectively engage and encourage the patient, making reliable data collection challenging^5^. Moreover, many are characterized by a high intra-patient variability (which might be caused by multiple factors e.g., activity days before the assessment, patient motivation), which limits the possibility to detect relevant changes in the patients’ trajectories. Lastly, the acquisition is time-consuming; it requires trained personnel and frequent visits to clinical sites. Alternatively, disease milestones such as Loss of Ambulation (LoA), Overhead Reach (OHR), and Hand to Mouth (HTM) have been used to identify modifiers of disease progression^6,7^. While informative, such milestones are not used as primary endpoints in clinical trials, as patients are typically enrolled early in life, and the duration of the study is too short to meet such milestones. Therefore, there is a need for alternative methods of assessment, such as blood biomarkers, which are objective since they are less dependent on patient cooperation, are quicker to collect, and can be studied regardless of the age of the patient and the duration of the study.

Multiple studies indicate that circulating proteins^8^ (e.g., myostatin), metabolites^9^ (e.g., creatine and creatinine), lipids^10^, miRNAs^11^, and mRNAs^12^ can discriminate patients from healthy controls, making them potential diagnostic markers. Additionally, recent reports indicate that serum protein levels are influenced by factors such as age^13,14^ and corticosteroid treatment^15,16^, highlighting the importance of examining biomarkers in relation to longitudinal functional performance and treatments. This study focuses particularly on two biomarkers, myostatin and the creatine-to-creatinine ratio (Cr/Crn), for the following reasons. Myostatin serum levels have been shown to be reduced in dystrophinopathy, e.g., DMD and Becker muscular dystrophy (BMD) patients^17,18^, while Cr/Crn was shown to be elevated^9,19^ with respect to healthy controls. Furthermore, both biomarkers have been found to be cross-sectionally associated with DMD clinical performance^9,17^, and associated with disease progression measured by timed tests and clinical scales in patients with BMD^20^. These associations are likely due to the fact that both myostatin levels and the conversion of creatine into creatinine depend on the amount of muscle tissue^21,22^. Myostatin is a negative regulator of muscle growth, primarily expressed in muscle tissue^23^. Creatine, synthesized by the liver and used as energy buffer during muscular contractions, is non-enzymatically metabolized in skeletal muscle into creatinine, which is then cleared by the kidneys^24^. While creatine and creatinine alone are confounded by diet and kidney function, calculating their ratio between these metabolites can be used to study the reduced conversion of creatine into creatinine. Such reduction can be visualized as an increase in Cr/Crn as muscle mass is lost.

For all these reasons, we hypothesized that myostatin and Cr/Crn could be associated with longitudinal clinical performance in DMD patients. Furthermore, we aimed to determine whether their limited variability, independent of patient motivation, makes them a valuable endpoint for future therapeutic and clinical trials. To address these hypotheses, we investigated serum concentrations of these biomarkers in a retrospective longitudinal cohort of DMD patients followed up at Leiden University Medical Center (LUMC). We associated these with functional tests and clinical milestones obtained during outpatient visits.

## Methods

### Data collection

Data were collected from DMD patients followed up at LUMC through the national disease registry for dystrophinopathies, the Dutch Dystrophinopathy Database^25^. Inclusion criteria were solely based on the confirmed genetic diagnosis of DMD, allowing patients to enter the study at different disease stages. Written informed consent was obtained from all participants and/or their legal representatives according to protocol B22.013 at LUMC. Clinical data and serum samples were collected at the same clinic and on the same visit.

Patients participated in yearly visits at the outpatient clinic, during which motor function tests, i.e., 6MWT, NSAA, 10MWT, and PUL2.0, were administered by a trained pediatric physiotherapist. Whenever the test was not performed due to factors such as the patient’s unwillingness, young age, or planning issues, missing values were registered. For 6MWT, NSAA, and 10MWT, the value was set to zero at the first non-ambulant visit, and to missing afterward.

Clinical milestones such as LoA, OHR, and HTM were also recorded during the study. LoA was reported by patients and/or caregivers and defined as the moment the individual was first unable to walk 5 meters unaided at home. In the absence of such a report, LoA was determined by the clinician as the inability to perform the 10MWT without support. In contrast, OHR and HTM were primarily assessed using the PUL2.0 and the Brooke and Vignos scores.

Data included the patient’s age at the time of the visit, body mass index (BMI), and information regarding the corticosteroid used. The treatment variable indicates whether or not the patient was taking corticosteroids (CS) at each specific visit (yes/no). Patients in the study were treated with different corticosteroids (prednisone, deflazacort, or vamorolone). Given the retrospective nature of the study, changes in CS type and regimen (daily or intermittent) were present over time for individual patients.

### Blood sampling

Serum samples were collected and prepared according to standard phlebotomy procedures. Samples at LUMC were left to clot for ~30 minutes, followed by 10 minutes of centrifugation at 2350g. Sample aliquots were frozen at –20°C for 1-2 months and then transferred to –80°C for long-term storage.

Creatine and creatinine were quantified by UPLC-MS/MS^26^. Stable isotope-labeled internal standards were added to 50 µl of serum. 500 µl of acetonitrile was added while vortexing, followed by centrifugation for 10 min at 12,000 x g. The supernatant was taken to dryness under a nitrogen flow, and the residue was reconstituted in 100 µl of water with 0.01% heptafluorobutyric acid (HFBA). 10 µl of the final solution was injected into a UPLC-MS/MS (Acquity Xevo TQ-XS, Waters, Milford, Massachusetts, USA) operating in positive ESI mode using MRMs for the preselected analytes. Separation was achieved on a BEH-C18 column (100 × 2.1 mm, 1.7 µm) using a linear gradient between solution B (acetonitrile/water, 4:1, v/v) and solution A (0.1% HFBA). The gradient (0.5 ml/min) was as follows: 0-2 min 95% A, 2-3 min 0% A, 3-3.1 min 95% A, and 3.1-5 min 95% A for re-equilibration. All gradient transitions were linear, and the total analysis time, including equilibration, was 5 min. Data processing was performed using MassLynx software (v4.2, Waters, Milford, Massachusetts, USA).

Myostatin was analyzed using a Quantikine ELISA kit (Bio-Techne #DGDF-80) in combination with a sample activation kit (BioTechne #DY010) and a quality control set 923 (BioTechne #QC98). In brief, 30 µl of serum was activated and subsequently processed and measured in duplicate according to Bio-Techne protocols using an ID3 plate reader at 450nm, with a correcting wavelength at 570nm^20^. Concentrations in pg/ml were calculated after averaging and interpolating the log-transformed values with a Four Parameter Logistic (4PL) regression. The coefficient of variation (CV) did not exceed 15,3% and had a mean of 3,7%.

### Statistical analysis

All the analyses were conducted using log2-transformed biomarkers. The log2 transformation addresses skewness and helps in the generation of distributions that are better suited to the assumptions of the analysis. Two linear mixed-effects models^27^ (LMM) were estimated to assess the association of log2-myostatin and log2-Cr/Crn ratio with age (centered around its mean) and CS treatment (treated/untreated), including patient-specific random effects. To evaluate the correlation between the two log2-biomarkers while accounting for the longitudinal structure of the data, we used the fitted values from the respective models and computed Spearman correlations^28^. The correlation was estimated at the median age, where the data were most densely concentrated, and stratified by CS treatment.

To determine the association between patients’ function and serum myostatin and Cr/Crn, we employed LMM with NSAA, 10MWT, 6MWT, and PUL2.0 as outcome variables and with patient-specific random intercepts. The number of available points was insufficient to include a random slope. As time-dependent predictors, we considered age, CS treatment (treated/untreated), and body mass index (BMI), as these may affect both patients’ performance^29^ and biomarkers’ levels^15^. Myostatin and Cr/Crn were used as predictors alone, as well as in combination. Zero values in the functional scores were maintained to capture the patients’ trajectory up to the loss of ambulation.

To examine the possible association of biomarkers with clinical milestones LoA, OHR, and HTM, we used time-dependent Cox models^30^. We included CS treatment and BMI. Significance of associations was evaluated for both biomarkers individually and jointly, with a p-value < 0.05.

Before fitting the models, log2-biomarkers, BMI, and age were also centered. Centering guarantees that the intercept represents the mean value of the outcome when all the continuous predictors are at their mean values, making the interpretation more reasonable.

Post-hoc sample size calculation based on functional test outcomes and log2-biomarkers was performed. For the calculation, an 80% power was assumed, and the effect size was defined using the Minimal Clinically Important Difference (MCID) quantities as reported in the literature for all outcomes^31–33^. To ensure that outcomes and biomarkers were comparable, we standardized (subtracting the mean and dividing by the standard deviation) the functional tests, log2-myostatin, and log2-Cr/Crn. We utilized the variance estimate obtained from the LMMs implemented on the outcomes and log2-biomarkers, with age and CS treatment as predictors and subject-specific random intercepts. The observed variance depended only on the within-patient variation. For more details on the power analysis, consult Section S1.

### Data availability

All data analyzed during this study have been anonymized and included as a supplementary file.

## Results

### Cohort characteristics

Data were collected from 74 DMD patients aged 4 to 24 years (median: 11.50, SD: 4.15) followed for up to 11 years (min: 1, max: 11, median: 5). One patient was considered an outlier and was removed from the analysis. This decision was made based on the trajectories of both myostatin and Cr/Crn. Even though the clinical results were in line with the rest of the patients’ group, the values of the biomarkers were largely outside the Interquartile Ranges (IQR). For comparison, the analysis results with the outlier are shown in Section S2. The remaining 73 patients had a total of 403 visits. Myostatin was quantified in 355 samples, and Cr/Crn in 368 samples. In 35 serum samples, both biomarkers could not be detected due to insufficient sample volume for analysis; for an extra 13 samples only Cr/Crn could be measured. The number of patients and serum samples used in each analysis varies due to missing values in the clinical data, as detailed for each analysis.

Section S3 includes visual timelines of patients’ functional test performances and illustrations showing how the likelihood of losing ambulation changes with age. Baseline characteristics are provided in Table 1. Since patients entered the study at various ages, the baseline was defined as each patient’s first visit. No patients entered the study unable to bring their hand to the mouth, whereas 9 patients could not reach overhead, and 14 were no longer ambulant. Most patients were untreated at baseline; 21 were receiving corticosteroids.

**Table 1.** Patients’ characteristics at baseline.

|  |  |
| --- | --- |
| **Age, years, median (min - max)** | 7 (4.10 - 20.10) |
| **CS treatment, n:** |  |
| • Treated/corticosteroids yes | 21 |
| • Untreated/corticosteroids no | 5 |
| **BMI, median (min - max)** | 16.70 (11.40 - 32.37) |
| **Myostatin, $\mu\text{mol/l}$, median (min - max)** | 1004 (248 - 3278) |
| **Creatine, $\mu\text{mol/l}$, median (min - max)** | 102.1 (65.9 - 158.4) |
| **Creatinine, $\mu\text{mol/l}$, median (min - max)** | 16.30 (6.27 - 31.85) |
| **Creatine/Creatinine ratio, median (min - max)** | 6.04 (2.69 - 16.14) |
| **10MWT, m/s, median (min - max)** | 1.78 (0 - 5.26) |
| **6MWT, m, median (min - max)** | 347 (0 - 525) |
| **NSAA, points, median (min - max)** | 23 (0 - 34) |
| **PUL2.0, points, median (min - max)** | 40 (30 - 42) |
| **Loss of ambulation, n** | 14 |
| **Loss of overhead reach, n** | 9 |
| **Loss hand to mouth, n** | 0 |
| *CS = corticosteroids, BMI = body mass index, 6MWT = six-minute walk test, NSAA = North Star Ambulatory Assessment, 10MWT = 10-meter walk-run test velocity, PUL2.0 = Performance of Upper Limb.* |  |

During the study, three patients received daily CS treatment, while 64 were on intermittent regimen. The remaining 6 never started CS treatment. The mean age of starting corticosteroids was 5.6 years. Overall, the most frequent corticosteroid used by the patients was prednisone (182 serum samples obtained in patients treated with the drug), followed by deflazacort (148 serum samples). Models of the functional scores, including only age, CS treatment, and BMI, showed that CS treatment was significantly associated with better function (10MWT: p-value = 0.006; 6MWT: p-value = 0.004, NSAA: p-value = 0.011, PUL2.0: p-value < 0.001), as detailed in Section S4.

The median age of LoA was 11.00 years, followed by OHR (13.3 years) and HTM (16.8 years). By the end of the study, 50 patients had lost ambulation, 41 had lost the ability to perform OHR, and 21 had lost HTM. Models excluding biomarkers showed that CS treatment significantly reduced the risk of LoA and OHR (Section S4).

### Association of biomarkers with age and CS treatment

The association with age was statistically significant for both myostatin and Cr/Crn (Section S5). Within the same treatment group, the model results estimated that (on average) log2-myostatin levels decrease by 0.10 units (p-value < 0.001), while log2-Cr/Crn values increase by 0.09 units (p-value < 0.001) for each year of age (Figure 1).

**Figure 1.**
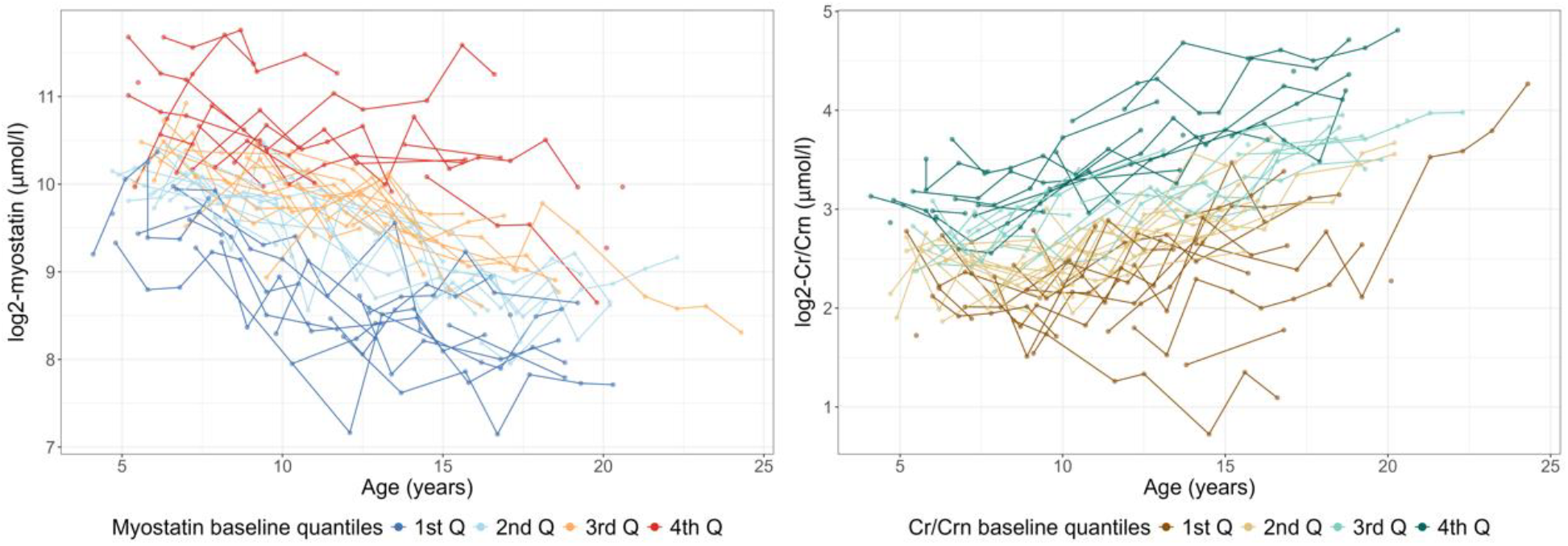
Longitudinal trajectories of myostatin (left) and creatinine/creatinine ratio (right) over the ages of the DMD patients. The patients’ profiles are colored based on the baseline quantile values of each biomarker. As baseline age varies across patients, the quartiles were defined using the random intercepts from linear mixed-effects models fitted to each biomarker. These intercepts can be interpreted as the estimated biomarker values at age zero. Cr/Crn = creatine/creatinine ratio, 1st Q = 1^st^ quartile.

Log2-myostatin was 0.46 units higher (p-value < 0.001) in treated versus untreated patients, while log2-Cr/Crn was 0.34 units lower (p-value < 0.001).

The correlation between myostatin and Cr/Crn was consistent across all ages. At the median age of 11.50 years, we observed a strong and negative correlation (R = –0.754, p < 0.001), with no evidence of differences when stratified by treatment group.

### Association of biomarkers with disease progression

To illustrate the strength and significance of the associations, we focused on model results from two functional tests: the NSAA for the lower limb function and the PUL2.0 for the upper limb function. Model estimates for the other functional tests are provided in Section S6. Models for NSAA and either myostatin, Cr/Crn, or both biomarkers estimated a yearly decline of 2.24-2.80 points (p-values < 0.001). Patients with a higher BMI tended to perform worse on the NSAA, with an average decrease of 0.80 points per unit increase in BMI (p-value < 0.001). CS treatment did not show consistent and statistically significant effects across the models. Both biomarkers were significantly associated with performance. Specifically, a unit decrease in log2-myostatin was associated with an average 4.73-point drop in NSAA (1st column, Table 2). In contrast, a unit increase in the log2-Cr/Crn corresponded to an average 11.40-point decrease in NSAA (2nd column, Table 2). However, when assessing the joint effect of biomarkers, only Cr/Crn remained statistically significant, and the effect size of myostatin diminished (3rd column, Table 2).

**Table 2.** Comparison of regression coefficients of the covariates of three different models for NSAA (left part) and PUL2.0 (right part): one with myostatin alone, one with Cr/Crn ratio alone, and one with both. The numbers of samples and patients used for the estimation of each model are reported as well. Cr/Crn = creatine/creatinine ratio, NSAA = North Star Ambulatory Assessment, PUL2.0 = Performance of Upper Limb, BMI = body mass index.

|  | NSAA |  |  |  |  |  | PUL2.0 |  |  |  |  |  |
| --- | --- | --- | --- | --- | --- | --- | --- | --- | --- | --- | --- | --- |
|  | Only Myostatin |  | Only Cr/Crn ratio |  | Both biomarkers |  | Only Myostatin |  | Only Cr/Crn ratio |  | Both biomarkers |  |
|  | Estimates | p-value | Estimates | P-values | Estimates | P-values | Estimates | p-value | Estimates | P-values | Estimates | P-values |
| Intercept | 4.85 | 0.037 | 5.22 | 0.007 | 6.09 | 0.003 | 25.50 | <0.001 | 25.52 | <0.001 | 26.00 | <0.001 |
| Age (years) | -2.80 | <0.001 | -2.28 | <0.001 | -2.24 | <0.001 | -1.77 | <0.001 | -1.32 | <0.001 | -1.25 | <0.001 |
| Treatment (yes) | 0.95 | 0.644 | 0.38 | 0.834 | -0.55 | 0.771 | 5.36 | <0.001 | 4.93 | <0.001 | 4.25 | <0.001 |
| BMI | -0.86 | <0.001 | -0.80 | <0.001 | -0.75 | <0.001 | -0.24 | 0.037 | -0.24 | 0.020 | -0.18 | 0.065 |
| Myostatin (log2) | 4.73 | <0.001 |  |  | 1.83 | 0.129 | 3.40 | <0.001 |  |  | 2.11 | 0.006 |
| Cr/Crn ratio (log2) |  |  | -11.40 | <0.001 | -10.02 | <0.001 |  |  | -7.18 | <0.001 | -6.22 | <0.001 |
| Observations | 186 |  | 193 |  | 186 |  | 183 |  | 183 |  | 183 |  |
| Patients | 58 |  | 62 |  | 58 |  | 55 |  | 55 |  | 55 |  |

The other two lower limb functional scores (6MWT and 10MWT) revealed similar results, supporting this pattern (Section S6). Disease severity increased with age and BMI, meaning that older patients or those with higher BMI on average scored lower on 6MWT and 10MWT. No statistically significant differences in progression were found between treated and untreated patients. There was a statistically significant positive association between myostatin levels and both 6MWT and 10MWT (p-values < 0.001), suggesting that performance declined with decreasing myostatin levels. The opposite association was found with the Cr/Crn ratio, showing that an increase in Cr/Crn corresponded to a deterioration of the performance (p-values < 0.001) (Figure 2).

**Figure 2.**
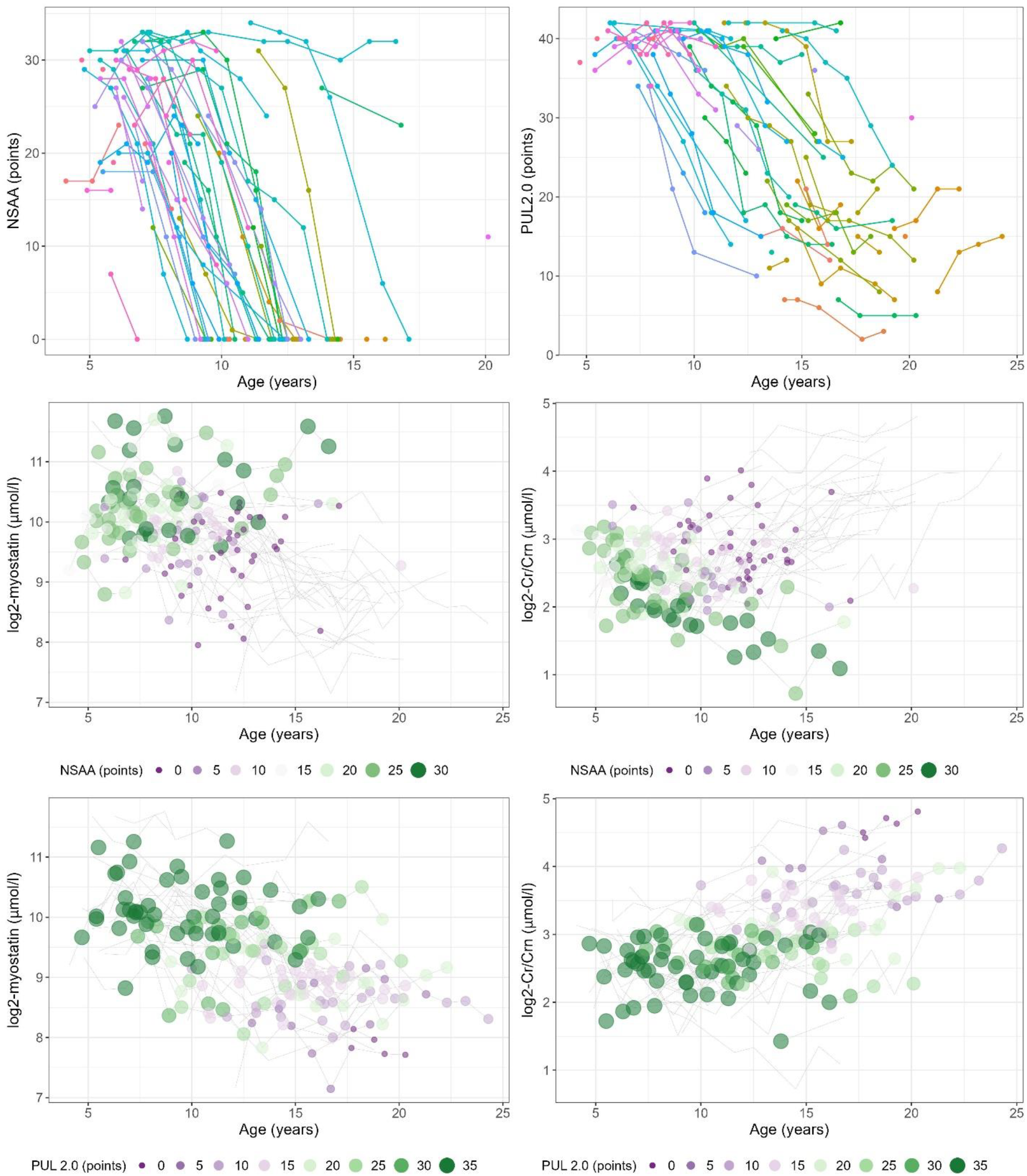
Longitudinal trajectory plots showing the NSAA and the PUL 2.0 scores progressions (first row) with age for all patients. Each line, distinguished by color, represents an individual patient. In the lower panels, trajectories illustrate how myostatin and Cr/Crn levels relate to NSAA (second row) and PUL 2.0 (third row) scores over time. Each gray line represents an individual patient’s biomarker trajectory, and each point corresponds to a clinical visit. Point color and size encode the functional score at each visit: larger, greener points indicate higher NSAA or PUL 2.0 values, while smaller, more violet points correspond to lower scores. NSAA = North Star Ambulatory Assessment, PUL2.0 = Performance of Upper Limb.

Similar conclusions were also obtained by modeling PUL2.0 trajectories (Table 2). For PUL2.0, treatment with CS retained a statistically significant effect in all models.

### Association of biomarkers with clinical milestones

We seek to assess whether myostatin and Cr/Crn were associated with disease milestones, namely LoA, OHR, and HTM (Figure 3).

**Figure 3.**
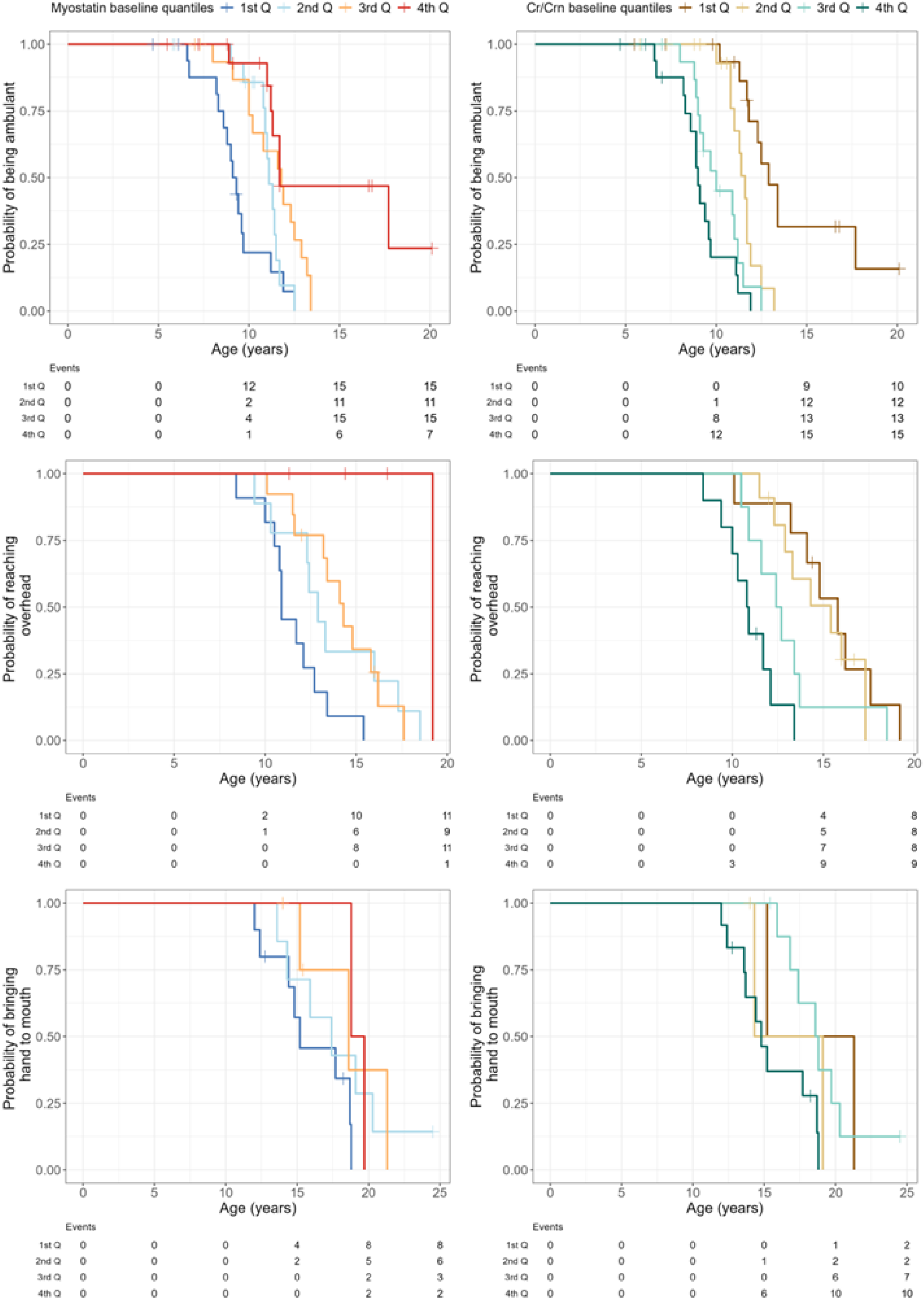
Kaplan-Meier plots showing the relationship between the clinical milestones and Myostatin (left) and Cr/Crn (right). Since baseline is different for each patient, we defined the baseline quartiles based on the random intercepts of the biomarkers’ linear mixed models. The cumulative number of events (LoA, OHR, or HTM) for each age is also reported. Cr/Crn = Creatine/Creatinine ratio, 1^st^ Q = 1^st^ quartile.

Survival models showed that those with lower log2-myostatin levels were more likely to lose ambulation within the following year (Table 3). Specifically, patients with a one-unit lower log2-myostatin value had a 42% (1 – 0.58) higher likelihood of losing ambulation, reflecting a more rapid decline in motor function compared to those with higher log2-myostatin levels. Conversely, patients with one unit lower values of log2-Cr/Crn were more likely to remain ambulant (Table 3). Subjects having higher log2-ratio experienced a more rapid decline in ambulation, with their hazard increasing approximately 3.67-fold. However, when evaluating the combined effect of both biomarkers, myostatin was no longer statistically significant, while the impact of Cr/Crn on the chance of losing ambulation increased (Table 3).

**Table 3.** Comparison of hazard ratios of the covariates of three different models for LoA: one with Myostatin alone, one with Cr/Crn ratio alone, and one with both.

|  | Only Myostatin |  | Only Cr/Crn ratio |  | Both biomarkers |  |
| --- | --- | --- | --- | --- | --- | --- |
|  | HR | P-values | HR | P-values | HR | P-values |
| CS Treatment (yes) | 0.36 | 0.025 | 0.56 | 0.205 | 0.53 | 0.173 |
| BMI | 1.03 | 0.318 | 1.06 | 0.100 | 1.06 | 0.086 |
| Log2-myostatin ($\mu\text{mol/l}$) | 0.58 | 0.003 | | | 1.40 | 0.234 |
| Log2-Cr/Crn ($\mu\text{mol/l}$) | | | 3.67 | <0.001 | 5.00 | <0.001 |

CS = corticosteroids, BMI = Body mass index, HR = Hazard Ratios, Cr/Crn = creatine/creatinine ratio.

In time-to-event models for OHR and HTM, both biomarkers were significantly associated with the outcome, but only when the myostatin and Cr/Crn were analyzed separately (Section S7). BMI was not found to be associated with LoA, while CS treatment significantly decreased the likelihood of LoA only in the model that included myostatin.

### Post-hoc sample size calculation

We wondered whether using biomarkers as endpoints in a hypothetical clinical trial could reduce the required sample size while preserving statistical power. Previous studies in DMD identified a 0.21 m/s decline in 10MWT or a 3-point decline in NSAA over one year as MCID^32,33^. To detect these effects with 80% power, the required sample size varies depending on the chosen outcome measure and its variability. In this analysis, we estimated the number of subjects needed to detect MCIDs, accounting for the observed variability of functional tests and biomarkers. Standardized values were used to compare the power analyses based on functional tests or biomarkers. By transforming the standardized values back to the original scales, we were able to determine the changes in log2-myostatin and log2-Cr/Crn over one year corresponding to a 3-point decline in NSAA, a 0.21 m/s decline in 10MWT (Figure 4), and a 30-meter decline in the 6MWT (Section S1).

**Figure 4.**
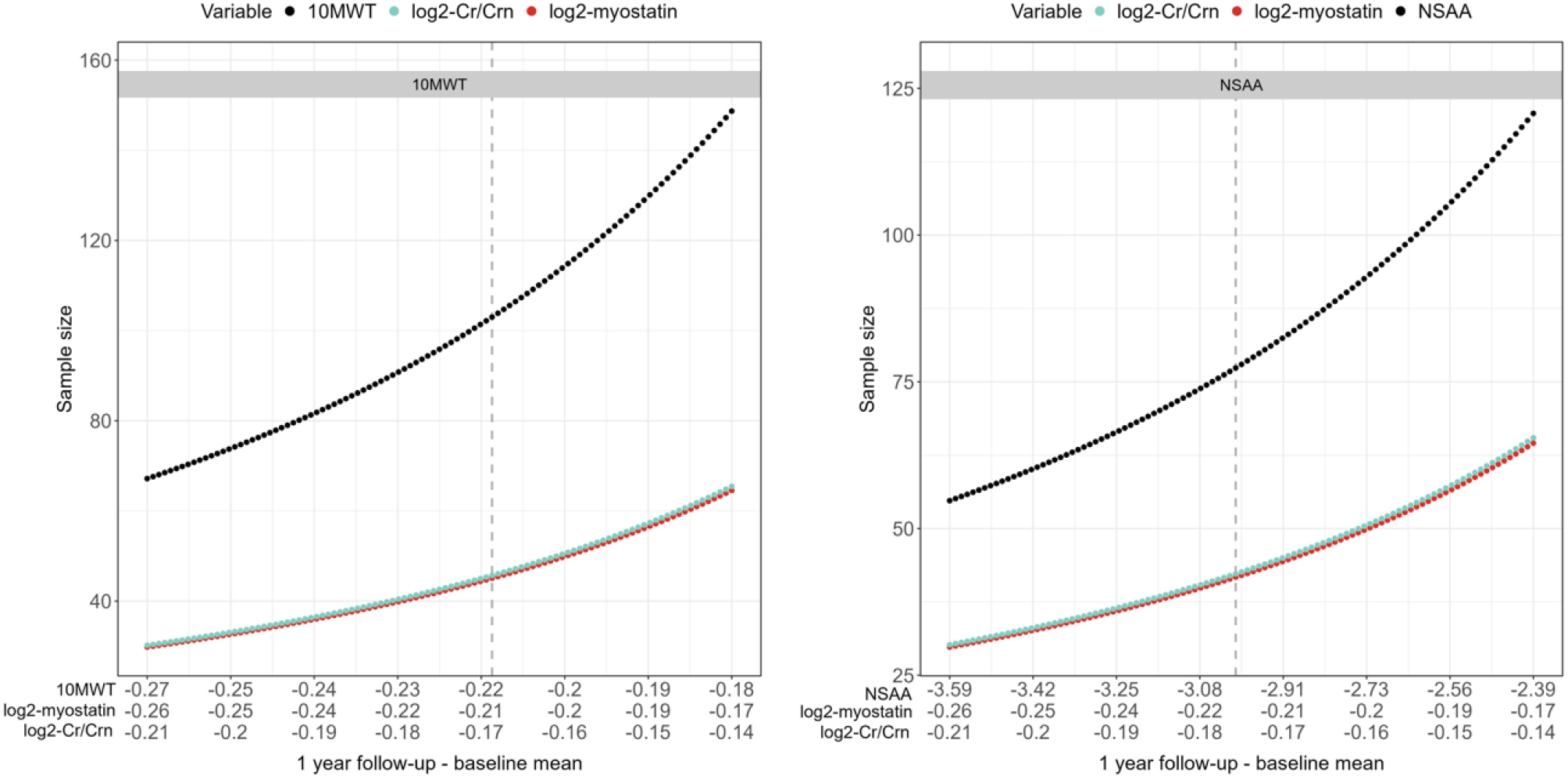
Number of DMD patients needed to detect an effect in one year since baseline in a 1:1 randomized trial. Comparison between the number of patients needed to detect the same effect in the 10MWT, in Myostatin, and Cr/Crn ratio (left); and the same effect in the NSAA, in Myostatin, and Cr/Crn ratio (right). Numbers on the x-axis are the effect sizes of the corresponding variables. The dashed vertical gray line indicates the point that corresponds, respectively, to the 0.21 m/s decline and the 3-point drop.

The observed standard deviations between one-year follow-up observations of the standardized NSAA and 10MWT were, respectively, 0.79 and 0.88. The within-patient variances of the standardized biomarkers were instead lower: 0.56 for log2-myostatin and 0.57 for log2-Cr/Crn ratio.

Power analysis estimated that almost 80 patients are needed to observe a 3-point NSAA decline over 1 year with 80% power in a 1:1 randomized trial, whereas around 50 patients are sufficient to observe proportional changes in log2-myostatin or log2-Cr/Crn levels (Figure 4). A similar trend was observed when comparing the sample sizes needed to detect equivalent effects in biomarkers versus the 10MWT. Using log2-myostatin and log2-Cr/Crn as endpoints significantly reduced the number of subjects needed: achieving 80% power to detect an effect equivalent to the MCID of the 10MWT in 1 year requires almost 2 times fewer patients compared to the 10MWT itself.

## Discussion

In this single-center, retrospective longitudinal study, we used real-world data to evaluate serum myostatin and the creatine-to-creatinine ratio as monitoring biomarkers in DMD. We showed that both biomarkers were affected by age, with myostatin progressively decreasing and Cr/Crn correspondingly increasing over time. We showed how these biomarkers were significantly associated with lower and upper limb functional tests and with lower and upper limb disease milestones. Importantly, incorporating these biomarkers as predictors of clinical function attenuated the statistical effect of corticosteroid treatment and eliminated its previously significant association with disease progression. Finally, we show that a hypothetical trial using Cr/Crn or myostatin as an endpoint would require fewer patients compared to the same trial where functional outcomes are used, due to the reduced intra-patient variation for these biomarkers compared to functional tests.

This study leveraged on a large retrospective cohort of 74 DMD patients followed at LUMC for up to 11 years. The long observational time allowed to confirm how Cr/Crn increases with age, as previously reported by Boca et al. (2016) in a cross-sectional study in 51 patients, and the decrease of myostatin as reported by Burch et al. (2017) in a cross-sectional comparison of 74 DMD patients. In the same study, Burch et al. (2017) collected longitudinal data for a short follow-up period (3.9 to 18.4 months) in a small sample of 26 DMD patients and could not reveal significant changes over that period. Since the number of follow-up samples was not reported, it is possible that the estimates were affected by the number of longitudinal observations or by the sensitivity of the different assays in our work compared to those of Burch et al.

Our longitudinal study confirmed the previously reported association of these biomarkers with disease severity. Previous cross-sectional studies showed that Cr/Crn and myostatin were associated with disease severity measures by functional tests such as the 6MWT, NSAA, and Forced Vital Capacity (FVC)^9,17^. We further showed that such associations extend for both biomarkers to the upper limb performance over multiple years. Importantly, in all studies, the directionality of the associations was confirmed, supporting the conclusion that decreasing myostatin and increasing Cr/Crn serum levels are associated with performance decline of lower and upper limb function.

The long follow-up also enabled the observation of disease milestones such as LoA, OHR, and HTM. While previous studies reported differences in these biomarkers between ambulant and non-ambulant patients^17^, we showed that both Cr/Crn and myostatin were significantly associated with the time to event, supporting the idea that they could be used to monitor how changes in these biomarkers are related to the likelihood of experiencing disease milestones.

Interestingly, inclusion of both myostatin and Cr/Crn showed that Cr/Crn ratio was a better predictor of function than myostatin. Results of the bivariate model were close to those obtained in the univariate Cr/Crn ratio model, with only small differences due to the effect of myostatin. This can be partly explained by the observed moderate correlation between myostatin and Cr/Crn ratio, justified by the likely relationship of these biomarkers with muscle mass. Such a strong correlation has already been reported in another longitudinal study in BMD patients^20^ (R = −0.85). Nonetheless, further studies in independent cohorts are needed to clarify the nature of their relationship (confounders, surrogates, etc.) and to validate our findings. Importantly, the myostatin concentrations measured in this study fall within the previously reported range (0.1–5 ng/ml). In contrast, previous reports of the Cr/Crn only partially align with our results, with all reported values being lower^9,19,20^ (<30 for the ratio, or <5 in log2 terms). The observed variability in Cr/Crn distribution may be attributed to differences in the methodology used. While previous studies used relative quantification methods^9,19^, in this study we used an absolute quantification method that provides a quantitative estimation of the levels of creatine and creatinine in serum (µmol/l).

Given the retrospective nature of the data, we corrected for confounding factors such as BMI and CS treatment, which are known to be associated with function and could affect the model estimates of the Cr/Crn and myostatin on patients’ performance. Our findings support the importance of maintaining a healthy weight for better functional performance. For instance, we observed a one-point increase in BMI corresponds to a decrease of 0.80 points in NSAA. However, BMI was not associated with the likelihood of clinical milestones, probably due to the low number of events. Notably, the association between BMI and function remained consistent even after including the biomarkers in the models, suggesting that Cr/Crn and myostatin do not capture differences in overall body weight.

Treatment with CS was significantly associated with better function in models without biomarkers, in line with extensive literature supporting the use of corticosteroids to maintain muscle function^29^. Interestingly, when Cr/Crn and myostatin were included in the models, the estimated effect of corticosteroid treatment was reduced and no longer statistically significant. The loss of significance may be due to the progressive reduction in the number of observations over time as patients lose ambulation, or to an insufficient number of events within each treatment group in the time-to-event analysis. The reduction in CS use estimates could also suggest that myostatin and Cr/Crn could partially capture the beneficial effects of treatment with steroids and function as potential surrogate. However, due to the observational and retrospective nature of the study, the interpretation of these associations should be carefully considered. The effect of CS is confounded by a variety of factors that were not controlled for in our study, such as the age at which treatment was first administered or the type or regimen of corticosteroid taken. Due to these and other factors (such as patients switching between CS medications over time, interruptions of treatment, and lean body mass, to mention a few), estimating such a relationship in prospective controlled studies such as the FOR-DMD^29^ would provide more reliable estimates. Importantly, our participants followed a 10 days on/10 days off corticosteroids regimen, but no information was available whether a patient was on the “on” or “off” phase of the CS treatment on the day of the visit/sampling. Additionally, blood sampling and functional assessments were not systematically obtained at the same time of the day, and we did not control for food intake. While more studies are required to confirm whether these biomarkers could eventually function as surrogate endpoints, it is noteworthy that the effect on functional tests of CS treatment could be captured by Cr/Crn and myostatin.

The data show how Cr/Crn and myostatin could be prospectively used in clinical trials to enrich the design and reduce patient inclusions. While patient inclusions are now based on functional performance mutation type (depending on the drug) and age ranges, new studies could include patients matching such clinical characteristics and prioritize patients with high Cr/Crn or low myostatin, as these patients represent likely decliners. Alternatively, including Cr/Crn and myostatin in the randomization process would allow a more fair comparison between treatment and placebo arms. Our post-hoc power analysis showed that using myostatin and Cr/Crn as endpoints can substantially reduce the number of patients needed to power a study. A hypothetical study with 1:1 randomization and 80% power, based on changes in myostatin or Cr/Crn, would halve the number of subjects needed compared to a study powered on clinical outcomes. This finding can be explained by the lower within-patient variability in biomarker levels compared to functional tests. This allows to have greater sensitivity in detecting changes across follow-up observations, compared to clinical outcomes. However, these conclusions are based on our observed data and require analysis in independent cohorts to assess whether these biomarkers could be informative during the randomization process or be used as primary, secondary, or exploratory endpoints.

In conclusion, we show that Cr/Crn and myostatin provide valuable information as monitoring biomarkers in DMD, as higher Cr/Crn and lower myostatin levels were associated with worse performance and higher risk of disease milestones such as loss of ambulation after adjusting for age, CS treatment, and BMI. Moreover, using these biomarkers as for patient stratification or as endpoints in future clinical trials may enhance such studies and contribute to reducing the number of subjects needed. Future studies with myostatin and Cr/Crn should aim to narrow the context of use of these biomarkers in DMD and ideally in other neuromuscular conditions, for which initial evidence has been recently reported^34–36^.

## Acknowledgements

This study was funded by Duchenne Parent Project NL (Project 22.010). The DDD and biobank have been supported by Spieren voor Spieren. The work was also partly supported by the National Institute Of Neurological Disorders And Stroke of the National Institutes of Health under Award Number # R61NS119639 (Co-I: Spitali, Tsonaka). The work was carried out within the framework of the European Reference Network for Neuromuscular Diseases (ERN EURO-NMD). The authors thank all participants and their families for contributing to the study.

## Author contributions

P.S., H.K., and E.H.N. contributed to the conception of the study and funding acquisition; N.I., P.S., M.N., and S.V. contributed to the acquisition of the data, and C.D. and R.T. to the analysis of the data. All the authors contributed to the drafting of the manuscript.

## References

1. Duan D, Goemans N, Takeda S, Mercuri E, Aartsma-Rus A. Duchenne muscular dystrophy. Nat Rev Dis Primers. 2021;7(1):1–19. doi:10.1038/s41572-021-00248-3

2. LBA02-09 EMBARK: A Phase 3 Randomized Study of Enzalutamide or Placebo Plus Leuprolide Acetate and Enzalutamide Monotherapy in High-risk Biochemically Recurrent Prostate Cancer. J Urol. 2023;210(1):224–226. doi:10.1097/JU.0000000000003518

3. Henzi BC, Schmidt S, Nagy S, et al. Safety and efficacy of tamoxifen in boys with Duchenne muscular dystrophy (TAMDMD): a multicentre, randomised, double-blind, placebo-controlled, phase 3 trial. The Lancet Neurology. 2023;22(10):890–899. doi:10.1016/S1474-4422(23)00285-5

4. Mercuri E, Vilchez JJ, Boespflug-Tanguy O, et al. Safety and efficacy of givinostat in boys with Duchenne muscular dystrophy (EPIDYS): a multicentre, randomised, double-blind, placebo-controlled, phase 3 trial. The Lancet Neurology. 2024;23(4):393–403. doi:10.1016/S1474-4422(24)00036-X

5. Role of motivation on performance of the 6-minute walk test in boys with Duchenne muscular dystrophy. Developmental Medicine & Child Neurology. 2015;57(S5):57–58. doi:10.1111/dmcn.94_12887

6. Bello L, Kesari A, Gordish-Dressman H, et al. Genetic modifiers of ambulation in the cooperative international Neuromuscular research group Duchenne natural history study. Annals of Neurology. 2015;77(4):684–696. doi:10.1002/ana.24370

7. Flanigan KM, Waldrop MA, Martin PT, et al. A genome-wide association analysis of loss of ambulation in dystrophinopathy patients suggests multiple candidate modifiers of disease severity. Eur J Hum Genet. 2023;31(6):663–673. doi:10.1038/s41431-023-01329-5

8. Hathout Y, Brody E, Clemens PR, et al. Large-scale serum protein biomarker discovery in Duchenne muscular dystrophy. Proc Natl Acad Sci U S A. 2015;112(23):7153–7158. doi:10.1073/pnas.1507719112

9. Spitali P, Hettne K, Tsonaka R, et al. Cross-sectional serum metabolomic study of multiple forms of muscular dystrophy. Journal of Cellular and Molecular Medicine. 2018;22(4):2442–2448. doi:10.1111/jcmm.13543

10. Srivastava NK, Pradhan S, Mittal B, Gowda GAN. High resolution NMR based analysis of serum lipids in Duchenne muscular dystrophy patients and its possible diagnostic significance. NMR Biomed. 2010;23(1):13–22. doi:10.1002/nbm.1419

11. Llano-Diez M, Ortez CI, Gay JA, et al. Digital PCR quantification of miR-30c and miR-181a as serum biomarkers for Duchenne muscular dystrophy. Neuromuscular Disorders. 2017;27(1):15–23. doi:10.1016/j.nmd.2016.11.003

12. Signorelli M, Ebrahimpoor M, Veth O, et al. Peripheral blood transcriptome profiling enables monitoring disease progression in dystrophic mice and patients. EMBO Molecular Medicine. 2021;13(4):e13328. doi:10.15252/emmm.202013328

13. Signorelli M, Ayoglu B, Johansson C, et al. Longitudinal serum biomarker screening identifies malate dehydrogenase 2 as candidate prognostic biomarker for Duchenne muscular dystrophy. Journal of Cachexia, Sarcopenia and Muscle. 2020;11(2):505–517. doi:10.1002/jcsm.12517

14. Strandberg K, Ayoglu B, Roos A, et al. Blood-derived biomarkers correlate with clinical progression in Duchenne muscular dystrophy. Journal of Neuromuscular Diseases. 2020;7(3):231. doi:10.3233/JND-190454

15. Hathout Y, Liang C, Ogundele M, et al. Disease-specific and glucocorticoid-responsive serum biomarkers for Duchenne Muscular Dystrophy. Sci Rep. 2019;9(1):12167. doi:10.1038/s41598-019-48548-9

16. Dang UJ, Ziemba M, Clemens PR, et al. Serum biomarkers associated with baseline clinical severity in young steroid-naïve Duchenne muscular dystrophy boys. Human Molecular Genetics. 2020;29(15):2481. doi:10.1093/hmg/ddaa132

17. Burch PM, Pogoryelova O, Palandra J, et al. Reduced serum myostatin concentrations associated with genetic muscle disease progression. J Neurol. 2017;264(3):541–553. doi:10.1007/s00415-016-8379-6

18. Mariot V, Joubert R, Hourdé C, et al. Downregulation of myostatin pathway in neuromuscular diseases may explain challenges of anti-myostatin therapeutic approaches. Nat Commun. 2017;8(1):1859. doi:10.1038/s41467-017-01486-4

19. Boca SM, Nishida M, Harris M, et al. Discovery of Metabolic Biomarkers for Duchenne Muscular Dystrophy within a Natural History Study. PLOS ONE. 2016;11(4):e0153461. doi:10.1371/journal.pone.0153461

20. Velde NM van de, Koeks Z, Signorelli M, et al. Longitudinal Assessment of Creatine Kinase, Creatine/Creatinineratio, and Myostatin as Monitoring Biomarkers in Becker Muscular Dystrophy. Neurology. 2023;100(9):e975–e984. doi:10.1212/WNL.0000000000201609

21. Wyss M, Kaddurah-Daouk R. Creatine and Creatinine Metabolism. Physiological Reviews. 2000;80(3):1107–1213. doi:10.1152/physrev.2000.80.3.1107

22. Mitra A, Qaisar R, Bose B, Sudheer SP. The elusive role of myostatin signaling for muscle regeneration and maintenance of muscle and bone homeostasis. Osteoporosis and Sarcopenia. 2023;9(1):1–7. doi:10.1016/j.afos.2023.03.008

23. Carnac G, Vernus B, Bonnieu A. Myostatin in the Pathophysiology of Skeletal Muscle. Current Genomics. 2007;8(7):415. doi:10.2174/138920207783591672

24. Brosnan JT, Brosnan ME. Creatine: Endogenous Metabolite, Dietary, and Therapeutic Supplement. Annual Review of Nutrition. 2007;27([Volume 27, 2007, Volume 27,]):241–261. doi:10.1146/annurev.nutr.27.061406.093621

25. van de Velde NM, Krom YD, Bongers J, et al. The Dutch Dystrophinopathy Database: A National Registry with Standardized Patient and Clinician Reported Real-World Data. J Neuromuscul Dis. 2024;11(5):1095–1109. doi:10.3233/JND-240061

26. Carling RS, Hogg SL, Wood TC, Calvin J. Simultaneous determination of guanidinoacetate, creatine and creatinine in urine and plasma by un-derivatized liquid chromatography-tandem mass spectrometry. Ann Clin Biochem. 2008;45(Pt 6):575–584. doi:10.1258/acb.2008.008029

27. Wu L. Mixed Effects Models for Complex Data. Chapman and Hall/CRC; 2009. doi:10.1201/9781420074086

28. Mukaka MM. A guide to appropriate use of Correlation coefficient in medical research. Malawi Medical Journal : The Journal of Medical Association of Malawi. 2012;24(3):69.

29. Guglieri M, Bushby K, McDermott MP, et al. Effect of Different Corticosteroid Dosing Regimens on Clinical Outcomes in Boys With Duchenne Muscular Dystrophy: A Randomized Clinical Trial. JAMA. 2022;327(15):1456–1468. doi:10.1001/jama.2022.4315

30. Therneau TM, Grambsch PM. Modeling Survival Data: Extending the Cox Model. Springer; 2000. doi:10.1007/978-1-4757-3294-8

31. McDonald CM, Henricson EK, Abresch RT, et al. The 6-minute walk test and other clinical endpoints in duchenne muscular dystrophy: Reliability, concurrent validity, and minimal clinically important differences from a multicenter study. Muscle & Nerve. 2013;48(3):357–368. doi:10.1002/mus.23905

32. Gupta VA, Pitchforth JM, Domingos J, et al. Determining minimal clinically important differences in the North Star Ambulatory Assessment (NSAA) for patients with Duchenne muscular dystrophy. PLOS ONE. 2023;18(4):e0283669. doi:10.1371/journal.pone.0283669

33. Duong T, Canbek J, Birkmeier M, et al. The Minimal Clinical Important Difference (MCID) in Annual Rate of Change of Timed Function Tests in Boys with DMD. J Neuromuscul Dis. 8(6):939–948. doi:10.3233/JND-210646

34. Moore U, Fernández-Simón E, Schiava M, et al. Myostatin and follistatin as monitoring and prognostic biomarkers in dysferlinopathy. Neuromuscular Disorders. 2023;33(2):199–207. doi:10.1016/j.nmd.2023.01.001

35. Tebbenkamp AT, Huggett SB, Lombardi V, et al. Protein biomarker signature in patients with spinal and bulbar muscular atrophy. JCI Insight. 2024;9(13):e176383. doi:10.1172/jci.insight.176383

36. de Albuquerque ALA, Chadanowicz JK, Giudicelli GC, et al. Serum myostatin as a candidate disease severity and progression biomarker of spinal muscular atrophy. Brain Communications. 2024;6(2):fcae062. doi:10.1093/braincomms/fcae062

